# How to minimize mechanical power during controlled mechanical ventilation

**DOI:** 10.1101/2024.11.05.24316778

**Authors:** Ben Fabry

**Affiliations:** Department of Physics, Friedrich-Alexander University Erlangen-Nürnberg, Germany

## Abstract

High intrapulmonary pressures, large tidal volumes, and elevated respiratory rates during controlled mechanical ventilation can lead to barotrauma, volutrauma, and atelectrauma. Mechanical power - defined as the product of the pressure-volume integral and respiratory rate - consolidates these three risk factors into a single, intuitive parameter. Several studies have demonstrated that higher mechanical power correlates with an increased risk of lung injury and mortality, prompting the suggestion that mechanical power should be minimized. However, under the constraint of maintaining a fixed alveolar minute ventilation and positive end-expiratory pressure (PEEP), it remains unclear how to adjust respiratory rate and tidal volume to minimize mechanical power. This study provides an analytical solution to this optimization problem. Accordingly, only the elastic component of mechanical power should be targeted for minimization. Regardless of lung elastance or resistance, or the mode and settings of the ventilator, the elastic power is minimized at a tidal volume equal to twice the anatomic dead space, or approximately 4.4 ml/kg of body weight.

## Introduction

Ventilator-induced lung injury (VILI) remains a major problem in the management of patients requiring controlled mechanical ventilation [1]. Risk factors such as high intrapulmonary pressures, large tidal volumes, and elevated respiratory rates can contribute to barotrauma, volutrauma, and atelectrauma of the lungs. To mitigate VILI, it is essential to optimize ventilator settings. Mechanical power (MP) has emerged as a comprehensive parameter to guide this optimization, as it integrates the risk factors contributing to VILI into a single, measurable, and intuitive value [2].

Mechanical power represents the total energy transferred from the ventilator to the respiratory system per unit time. It is calculated as the product of the mechanical work required to fill the lungs during a single inspiration (*WOB*) and the respiratory rate (*rr*). The mechanical work of breathing comprises three main components: (i) the work to overcome the elastic forces of the respiratory system (*WOB*_*el*_), (ii) the work to overcome resistive forces (*WOB*_*res*_), and (iii) the work to move air against the intrinsic positive end-expiratory pressure (*WOB*_*iPEEP*_).

Mechanical power increases with higher minute ventilation, increased elastance and resistance, and elevated lung opening pressure - all of which are linked to compromised lung function. Consequently, it is not surprising that higher mechanical power is correlated with more severe lung injury and higher mortality [1, 3]. Mechanical power should not be confused with a linear scale that reflects the risk of developing VILI in all patients equally [4]. Nonetheless, reducing mechanical power is generally considered beneficial in mitigating the risk of VILI [3, 5].

Mechanical power can be reduced by minimizing minute ventilation and positive end-expiratory pressure (PEEP), but not below clinically safe levels to avoid hypoventilation or atelectasis. Within the constraint of a fixed minute ventilation, mechanical power can be further reduced by adjusting the respiratory rate, tidal volume, and the inspiration-to-expiration ratio. This study explores how mechanical power varies with ventilator settings and identifies the optimal settings to minimize mechanical power.

## Methods

Formulating the mechanical power mathematically is most straightforward for traditional volume-controlled mechanical ventilation with constant inspiratory flow 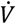 and an expiration phase in which airway pressure is maintained at a constant positive level (PEEP). For simplicity, we assume that the respiratory system behaves as a linear system similar to an elastic balloon with constant elastance *E*, which is the sum of lung and chest wall elastance, connected to the ventilator through a tube with constant resistance *R*, which is the sum of the airway resistance and the resistance of the endotracheal or tracheostomy tube. Then, 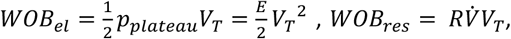 and *WOB*_*iPEEP*_ = *iPEEP V*_*T*_, with *V*_*T*_ being the tidal volume, and *p*_*plateau*_ being the end-inspiratory plateau pressure, which equals *E V*_*T*_.

The intrinsic PEEP is typically higher than the externally applied PEEP unless the expiratory time is much larger than the time constant *τ* for passive lung emptying, where *τ* = *R/E*. The intrinsic PEEP can be analytically computed by considering that the tidal volume *V*_*T*_ delivered during inspiration is only partially exhaled during the following expiration time *t*_*ex*_ so that the residual end-expiratory lung volume from that breath equals 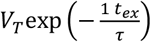. Additionally, there is a residual volume from two breaths prior, 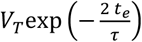, from three breaths prior,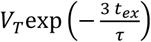 and so on, forming an infinite geometric series. Thus, the intrinsic PEEP is given by:

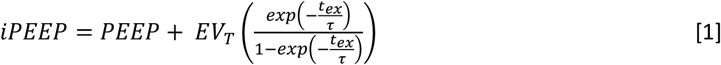

By combining all these factors, the total mechanical power can be computed using the following equation:

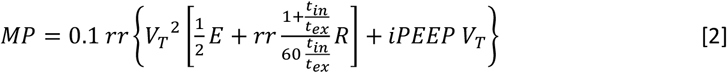

as derived, explained step by step, and validated in [1]. Here, *t*_*in*_ is the inspiration time, *V*_*T*_ is given in units of liters (L), *R* in mbar/L/s, *E* in mbar/L, rr in min^-1^, and the factor 0.1 converts the power into units of J/min.

Finding the optimal combination of *rr, V*_*T*_, and I:E ratio that minimizes mechanical power is not straight-forward. Eq. 1 contains several non-linear terms, and moreover *rr* and *V*_*T*_ cannot be independently set as they are constrained by the maintenance of a certain alveolar minute ventilation 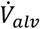, depending also on the anatomic dead space *V*_*D*_, according to

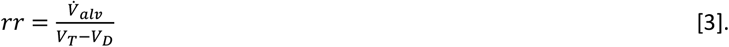

This introduces another non-linear term, in particular when the tidal volume approaches the anatomic dead space.

The optimal combination of *rr, V_T_*, and I:E ratio that minimizes mechanical power according to Eq. 2 can be found numerically. Although reducing PEEP will always reduce mechanical power, PEEP is usually adjusted so as to maximize lung recruitment and to prevent atelectasis, and is therefore assumed to be as low as clinically sensible. For simplicity, PEEP is set to zero for all subsequent calculations, as it does not affect the optimal combination of *rr, V_T_*, and I:E ratio that minimizes mechanical power. A Python script for performing the numerical power minimization can be downloaded from https://github.com/fabrylab/MP_ventilation.

## Results

All three components of mechanical power (elastic, resistive, and iPEEP) become very large when the tidal volume approaches the anatomic dead space (Fig. 1). This was first observed by Otis et al. [6] and is explained by the increase in total minute ventilation as a substantial fraction of the tidal volume is wasted on dead space ventilation. With increasing tidal volumes, the resistive power and the power associated with iPEEP decrease monotonically, whereas the elastic power first decreases but eventually increases (Fig. 1). The total mechanical power exhibits a minimum when the increase in elastic power exceeds the decrease in resistive and iPEEP power.

**Fig 1.**
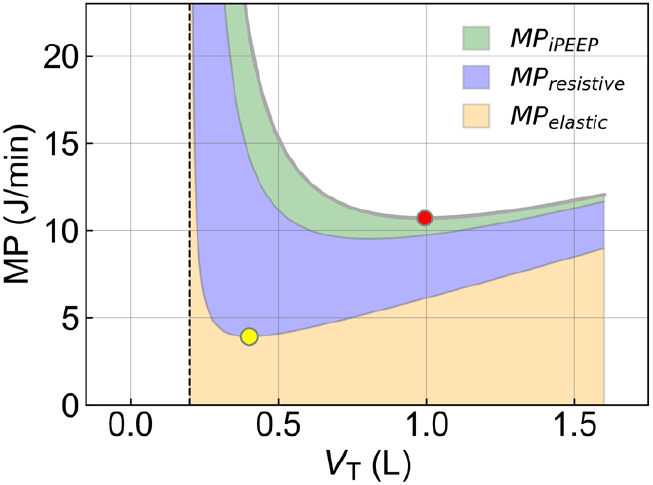
Components of mechanical power as a function of tidal volume. Resistive and iPEEP mechanical power monotonically decrease with increasing tidal volumes when alveolar minute ventilation is kept constant (here: 10 L/min), whereas elastic power first decreases until it reaches a minimum at 400 ml (yellow marker), which is twice the anatomic dead space (here: 200 ml, dashed vertical line), and then linearly increases. The total mechanical power reaches a minimum at a tidal volume of 993 ml (red marker). Elastance is set to 10 mbar/L (left), corresponding to a normal lung. Airway resistance is set to 1 mbar/L/s, and tube resistance is 7 mbar/L/s, corresponding to an 8 mm endotracheal tube [7].

Importantly, the total mechanical power exhibits a minimum at a considerably larger tidal volume than the minimum of the elastic power component (Fig. 1). In fact, for a range of clinically relevant patient parameters, the tidal volumes that minimize total mechanical power often substantially exceed the tidal volumes recommended in lung-protective ventilation protocols [8] (Fig. 2).

**Fig 2.**
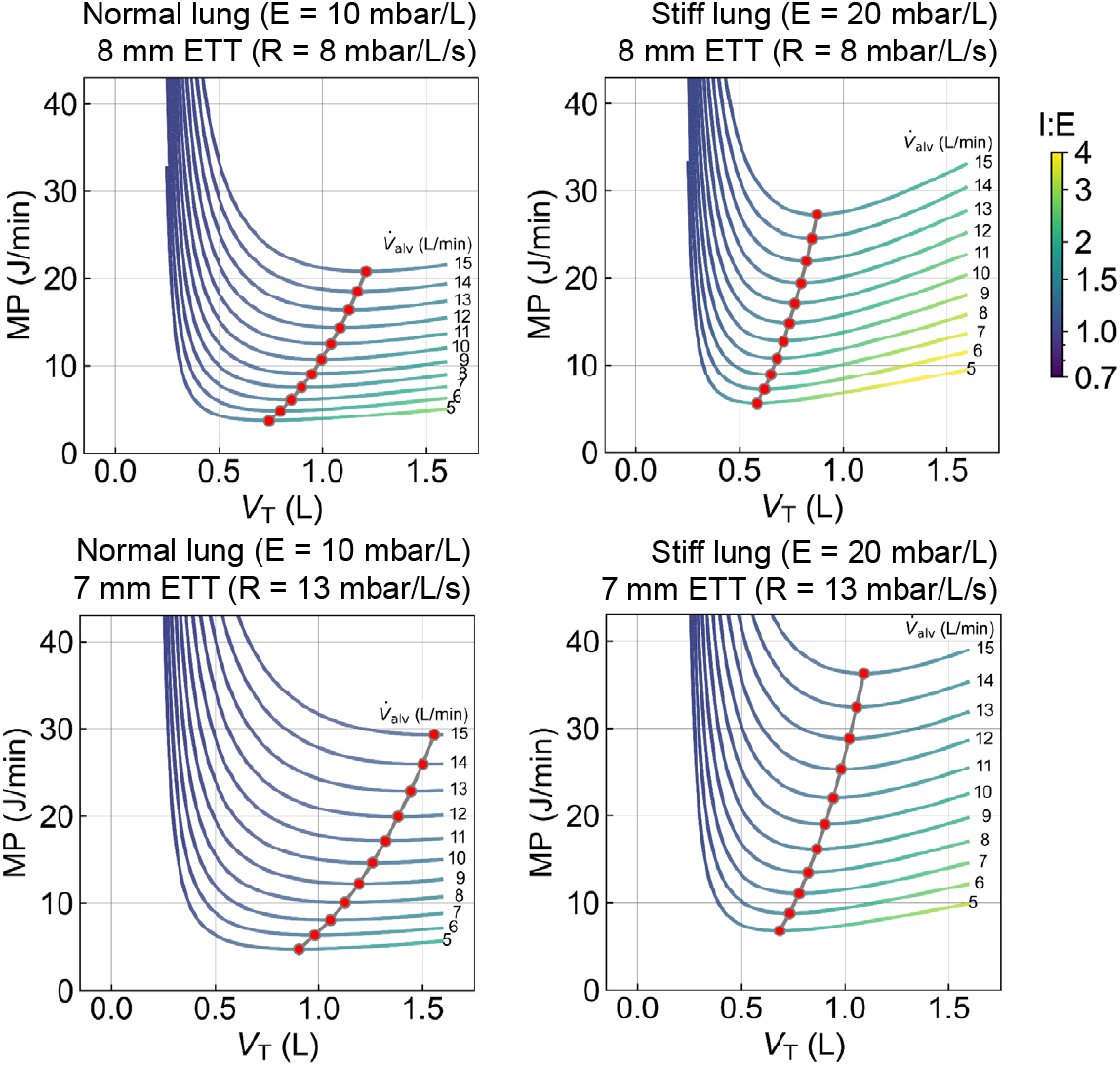
Mechanical power as a function of tidal volume. Each line represents a different alveolar minute ventilation. Line colors correspond to the I:E ratio that results in the lowest mechanical power. Elastance is set to 10 mbar/L (left), corresponding to a normal lung, or 20 mbar/L (right), corresponding to a stiff lung seen in mild to moderate ARDS. Airway resistance is set to 1 mbar/L/s, and tube resistance is either 7 mbar/L/s, corresponding to an 8 mm endotracheal tube (top), or 12 mbar/L/s, corresponding to a 7 mm endotracheal tube (bottom) [7]. Anatomic dead space is set to 200 mL. For each condition, the minimum of the MP versus V_T_ relationship is indicated by red symbols.

The observation that the total mechanical power exhibits a minimum only at large tidal volumes is attributable to the behavior of the resistive and iPEEP power (Fig. 1). The resistive power in most patients, unless they suffer from COPD or asthma, is dominated by the resistance of the endotracheal tube [7]. The endotracheal tube, however, is a component of the ventilator, not the patient. Whether a lower or higher mechanical power is consumed by the endotracheal tube is of no consequence for a patient under controlled mechanical ventilation, apart from the effect that the tube resistance has on intrinsic PEEP build-up. Resistive power consumption by the endotracheal tube does not damage the patient’s lungs. Similarly, the power consumed by the patient’s airway resistance, as far as we know, also does not damage the patient’s lungs.

It has been argued that high flow rates, which tend to increase resistive power, may potentially damage the lungs by causing higher strain rates at the tissue level [9]. But indirectly penalizing high strain rates through resistive power is arbitrary and obscures the physiological rationale behind the concept of power minimization. Therefore, for patients receiving controlled ventilation who are not spontaneously breathing, there is no plausible argument for including resistive power.

Similarly, the mechanical power associated with intrinsic PEEP, which like resistive power tends to decrease with higher tidal volumes, should not be included in a power minimization scheme either. Any build-up of intrinsic PEEP can be readily compensated by reducing the external PEEP by a corresponding amount – hence, there is no practical need to select a combination of low respiratory rate and large tidal volume just to reduce iPEEP build-up.

For these reasons, only the elastic component of mechanical power should be minimized. Then, Eq. 2 simplifies to

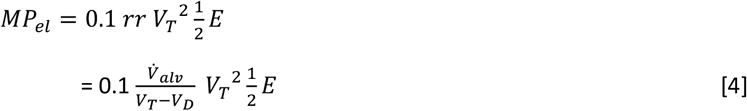

This approach has the added benefit that the elastic mechanical power is completely independent of the flow waveform, and hence Eq. 4 is applicable to both, pressure-controlled and volume-controlled modes of ventilation.

By taking the derivative of Eq. 4 with respect to *V*_*T*_ and setting it to zero, one finds that the elastic mechanical power is always minimized at a tidal volume that is exactly twice the anatomic dead space, regardless of respiratory elastance and alveolar minute ventilation. This holds true for the example shown in Fig. 1 as well.

In adults, the anatomic dead space of the lungs (in units of milliliters) scales approximately linearly with body weight *BW* (in units of kilograms), according to *V*_*D*_ ∼ 2.2 × *BW* [10]. Thus, a tidal volume of 4.4 ml/kg body weight minimizes elastic power. In ventilated patients, the dead space as estimated based on body weight needs to be corrected for the added dead space of external tubing or devices, and for the reduction in dead space due to intubation. Ideally, the anatomic dead space should be measured directly, for example, by capnography.

## Discussion

Numerous studies have demonstrated that high mechanical power of ventilation is correlated with a poor outcome [1-5]. Excessive mechanical power can be avoided by a sensible choice of minute ventilation and PEEP level, and – as shown here – can be further reduced by an optimal combination of tidal volume, respiratory rate, and I:E ratio. However, this study also finds that for a given alveolar ventilation, the total mechanical power reaches a minimum at low respiratory rates and large tidal volumes that are often beyond the range considered lung-protective. Moreover, around its minimum, the total mechanical power is insensitive to changes in tidal volume.

Two clinical recommendations can be drawn from this analysis. First, minimizing total mechanical power for a given alveolar minute ventilation can lead to potentially unsafe tidal volumes and should be discouraged. Second, because the curve of total mechanical power is relatively flat around its minimum over a wide range of tidal volumes, total mechanical power is an unsuitable guide for adjusting ventilator settings.

The shift of the total power minimum to large tidal volumes is rooted in the behavior of its individual components. The elastic power exhibits a minimum at a small tidal volume of twice the anatomic dead space, and then increases with tidal volume. By contrast, resistive and iPEEP power monotonically decrease with higher tidal volume. As a result, the total minimum shifts to large tidal volumes, especially in patients with low respiratory elastance and high resistance requiring high minute ventilation (Fig. 2).

As argued above, the resistive power is not known to cause lung damage. Moreover, the power associated with intrinsic PEEP buildup can be prevented by reducing the external PEEP. Hence, only the elastic power should be considered for optimizing the ventilator settings.

Focusing only on elastic power greatly simplifies the task of finding the optimal ventilator settings. Both for pressure-controlled or volume-controlled mechanical ventilation, the elastic power always reaches its minimum at exactly twice the anatomic dead space, regardless of minute ventilation, I:E ratio, or the respiratory parameters of the patient. This insight provides a straightforward approach for minimizing mechanical power:

1. Select a tidal volume that is twice the anatomic dead space (in adults, this corresponds to a tidal volume of approximately 4.4 ml/kg body weight).
2. Compute the respiratory rate that is needed to achieve the desired alveolar minute ventilation using Eq. 3.
3. Select the smallest external PEEP that prevents atelectasis and maintains open lungs.
4. Reduce the external PEEP by the amount of intrinsic PEEP buildup, as measured with an end-expiratory occlusion maneuver, or as calculated using Eq. 1.

The I:E ratio does not affect elastic power. However, an I:E ratio near unity may be recommended as it balances the peak inspiratory and expiratory flow, leading to the lowest overall strain rates.

This novel power minimization approach advocates tidal volumes at the lower end of the recommended range for lung-protective ventilation, but is not in conflict with current clinically established guidelines.

## Data Availability

All data produced in the present work are contained in the manuscript

https://github.com/fabrylab/MP_ventilation

